# The WORC database: MRI and CT scans, segmentations, and clinical labels for 930 patients from six radiomics studies

**DOI:** 10.1101/2021.08.19.21262238

**Authors:** Martijn P.A. Starmans, Milea J.M. Timbergen, Melissa Vos, Guillaume A. Padmos, Dirk J. Grünhagen, Cornelis Verhoef, Stefan Sleijfer, Geert J.L.H. van Leenders, Florian E. Buisman, Francois E.J.A. Willemssen, Bas Groot Koerkamp, Lindsay Angus, Astrid A.M. van der Veldt, Ana Rajicic, Arlette E. Odink, Michel Renckens, Michail Doukas, Rob A. de Man, Jan N.M. IJzermans, Razvan L. Miclea, Peter B. Vermeulen, Maarten G. Thomeer, Jacob J. Visser, Wiro J. Niessen, Stefan Klein

## Abstract

The WORC database consists in total of 930 patients composed of six datasets gathered at the Erasmus MC, consisting of patients with: 1) well-differentiated liposarcoma or lipoma (115 patients); 2) desmoid-type fibromatosis or extremity soft-tissue sarcomas (203 patients); 3) primary solid liver tumors, either malignant (hepatocellular carcinoma or intrahepatic cholangiocarcinoma) or benign (hepatocellular adenoma or focal nodular hyperplasia) (186 patients); 4) gastrointestinal stromal tumors (GISTs) and intra-abdominal gastrointestinal tumors radiologically resembling GISTs (246 patients); 5) colorectal liver metastases (77 patients); and 6) lung metastases of metastatic melanoma (103 patients). For each patient, either a magnetic resonance imaging (MRI) or computed tomography (CT) scan, collected from routine clinical care, one or multiple (semi-)automatic lesion segmentations, and ground truth labels from a gold standard (e.g., pathologically proven) are available. All datasets are multicenter imaging datasets, as patients referred to our institute often received imaging at their referring hospital. The dataset can be used to validate or develop radiomics methods, i.e., using machine or deep learning to relate the visual appearance to the ground truth labels, and automatic segmentation methods. See also the research article related to this dataset: Starmans et al., *Reproducible radiomics through automated machine learning validated on twelve clinical applications*, Submitted.

**Specifications Table:** 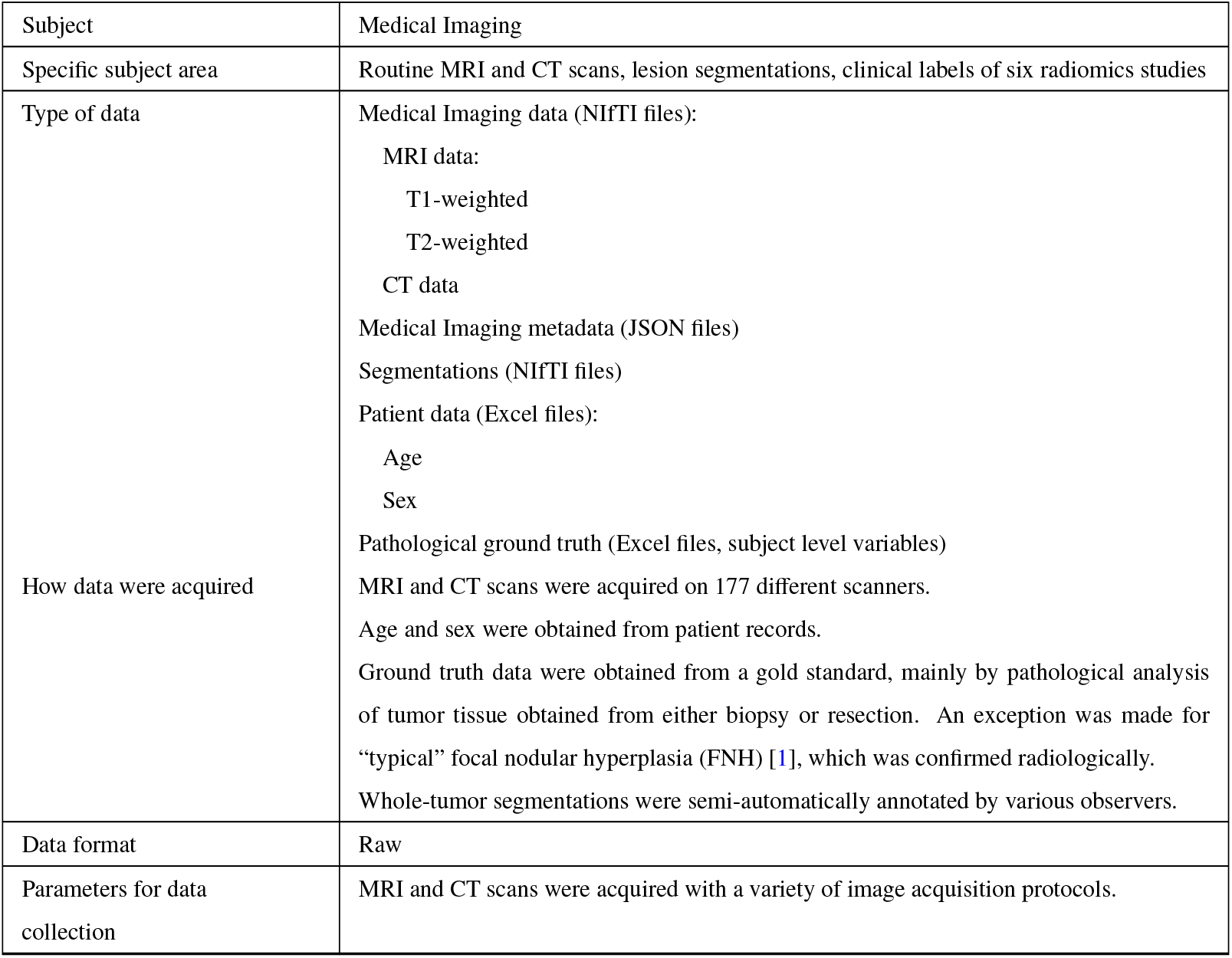

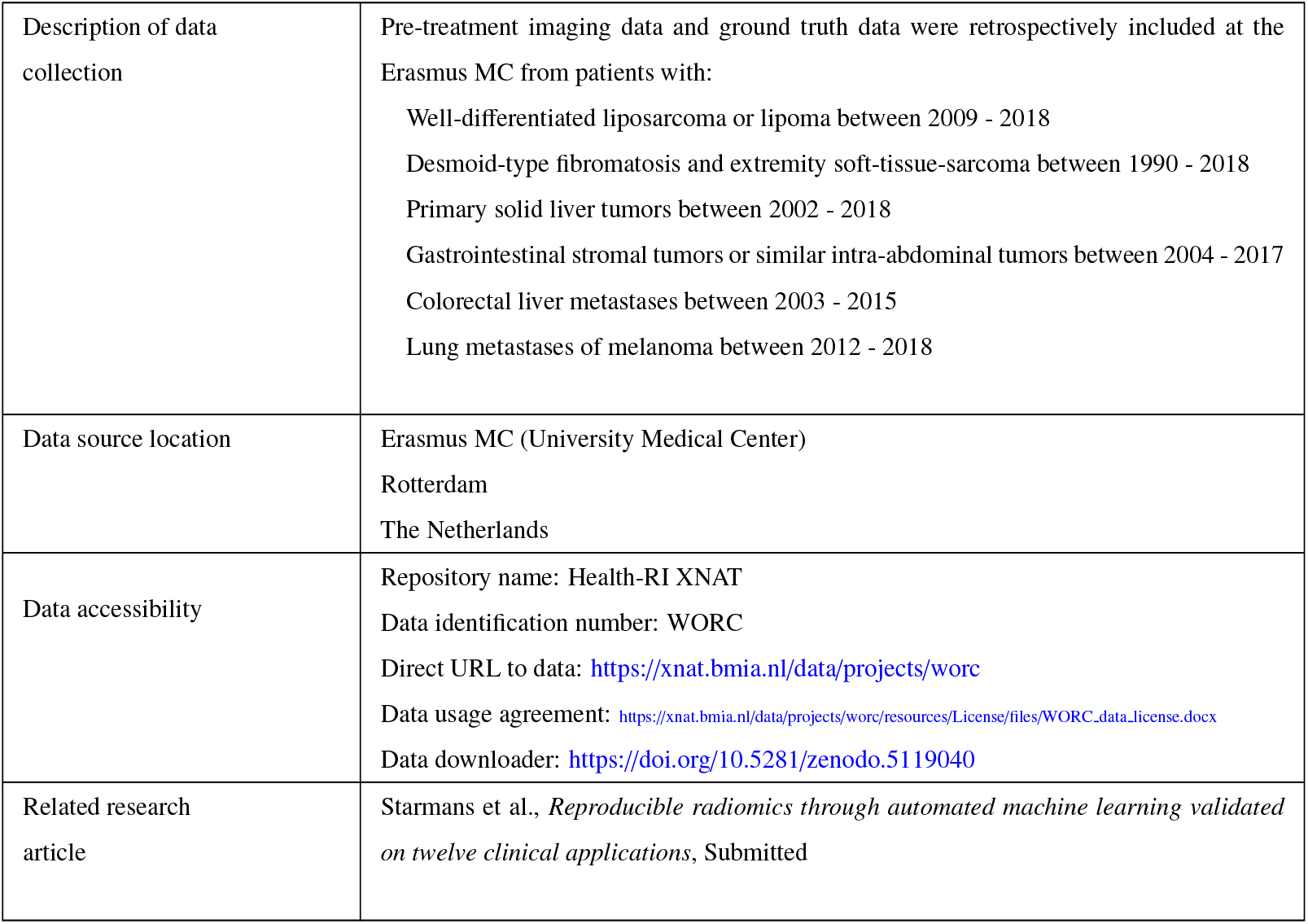

## 1. Value of the Data

- This dataset provides imaging data, outlined lesions, age, sex, and ground truth labels (e.g., diagnosis, genetic mutations, biological characteristics), mostly obtained from pathology, for a large number of patients from six different cancer studies. Publicly sharing imaging data with ground truth labels and segmentations benefits reproducibility, enables external validation, and hence accelerates transition to clinical practice [2, 3, 4]. This dataset has been collected in routine clinical care at multiple centers, thus representing the real-life variability and heterogeneity of the data. For these reasons, this dataset is a valuable resource.
- This dataset will be beneficial for researchers working on computer aided diagnosis for cancer based on imaging, specifically in the areas of liposarcoma, desmoid type-fibromatosis, gastrointestinal stromal lesions, sarcoma, primary liver cancer, (colorectal) liver metastases, and (melanoma) lung metastases.
- This data can be used to validate or develop radiomics methods (i.e., using conventional machine learning or deep learning to relate the visual appearance to the ground truth labels) and automated segmentation methods. For example, the data can be used as a large, heterogenous independent test set, or to increase the size and heterogeneity of train sets for developing new methods.

## 2. Data Description

The WORC dataset contains 930 patients and is composed of six radiomics studies, coined the Lipo (subsection 2.1), Desmoid (subsection 2.2), Liver (subsection 2.3), GIST (subsection 2.4), CRLM (subsection 2.5), and Melanoma (subsection 2.6) dataset. All datasets were collected at the Erasmus MC, Rotterdam, the Netherlands, but are multicenter imaging datasets, as patients referred to our institute often received imaging at their referring hospital. Example images of each dataset are shown in Figure 1.

**Figure 1:**
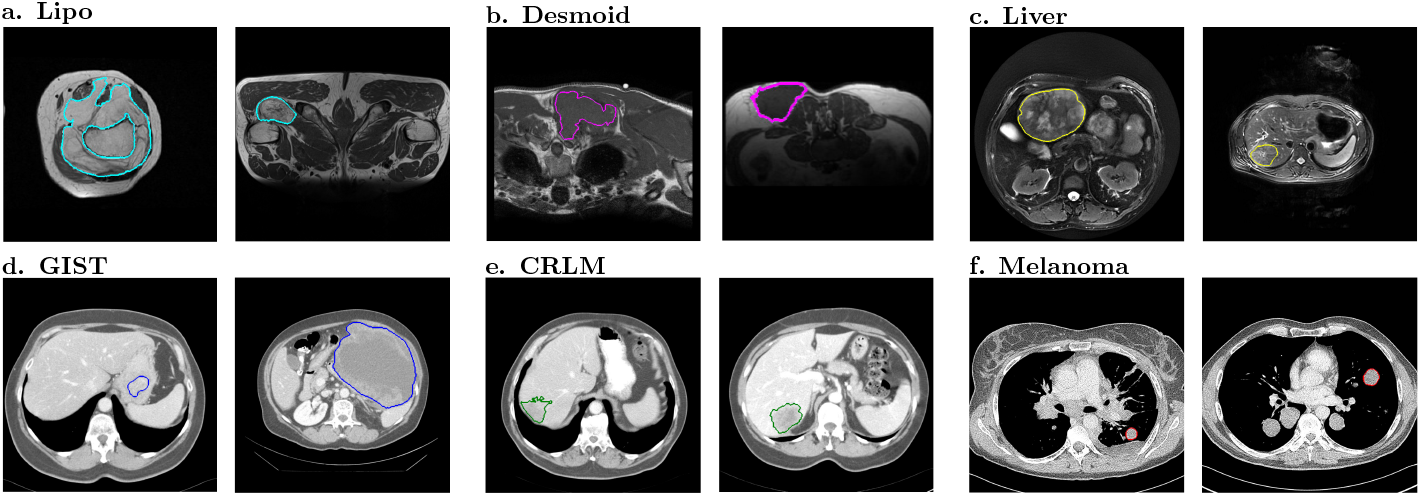
Examples of the 2D slices from the 3D imaging data from the six datasets included in the WORC dataset. For each dataset, for one patient of each of the two classes, the 2D slice in the primary scan direction (e.g., axial) with the largest area of the segmentation is depicted; the boundary of the segmentation is projected in color on the image. The datasets included were from different clinical applications: a. lipomatous tumors [9]; b. desmoid-type fibromatosis [10]; c. primary solid liver tumors [11]; d. gastrointestinal stromal tumors [12]; e. colorectal liver metastases [13]; and f. melanoma [14].

For each study, five different sources of data are provided:

1. Routine clinical MRI (Lipo, Desmoid, Liver) or CT (GIST, CRLM, Melanoma) scans
2. Details on the acquisition protocols (subsection 2.7)
3. Lesion segmentations
4. Age and sex
5. Pathological ground truth labels

The data is available on an XNAT server; an online platform to store (medical) imaging data in a standardized way, allowing access through both a Graphical User Interface (GUI) and an Application Programming Interface (API) [5]. The datasets for this study are publicly hosted on the Health-RI XNAT ^1^. Code to download the data locally, and code to reproduce the experiments from Starmans et al. [6] on these datasets, have been released open-source [7].

For each study, details on the ground truth labels and the data collection are given in the respective subsections. The acquisition protocol details for all studies are described in subsection 2.7. The scans have been converted from DICOM to NIfTI using the dcm2niix toolbox version v1.0.20180518 [8]. For each patient, a single scan is included and provided as NIfTI files named *“image*.*nii*.*gz”*. The associated details on the scan acquisition protocol are given in a JSON file named *“metadata*.*json”*. The corresponding segmentation is given in the NIfTI file *“segmentation*.*nii*.*gz”*, where a label of 1 indicates a lesion and a label of 0 indicates background. For the CRLM dataset, multiple segmentations of various lesions made by multiple observers are given, see subsection 2.5. The ground truth pathological labels for all studies are combined in the Excel sheet *“Clinical_data*.*xlsx”* and as labels on subject level in the `XNAT` project to allow for easier automatic processing.

### 2.1. The Lipo dataset

This dataset consists of 115 patients with either a well-differentiated liposarcoma (WDLPS) (*N* = 58) or lipoma (*N* = 58), as described in Vos et al. [9]. One patient has both a WDLPS and a lipoma, thus the dataset in total contains 116 lesions. For each patient, a T1-weighted MRI scan is provided. The ground truth label, i.e., whether a lesion was a WDLPS or lipoma, is represented by the *MDM2* amplification. The *MDM2* amplification status for each patient is provided, where patients have label 1 if the lesion was a WDLPS, and label 0 if the lesion was a lipoma.

For the patient with both a WDLPS and a lipoma, a segmentation is provided for each lesion: *“segmentation_WDLPS*.*nii*.*gz”* and *“segmentation_lipoma*.*nii*.*gz”*

### 2.2. The Desmoid dataset

This dataset consists of 203 patients with either desmoid-type fibromatosis (DTF) (*N* = 72) or extremity soft-tissue sarcomas (STS), i.e, the non-DTF group (*N* = 131), as described in Timbergen et al. [10]. The non-DTF group consists of 64 myxofibrosarcomas, 31 leiomyosarcomas, and 36 myxoid liposarcomas. For each patient, a T1-weighted MRI scan is provided. The ground truth label, i.e., whether a lesion was a DTF or one of the non-DTF phenotypes, was confirmed by histology. The differential diagnosis for each patient is provided, where patients have label 1 if the lesion was a DTF, and label 0 if the lesion was a non-DTF. The subtype of the non-DTF lesions is also provided.

### 2.3. The Liver dataset

This dataset consists of 186 patients with either a malignant (*N* = 94) or benign (*N* = 93) primary solid liver tumor, as described in Starmans et al. [11]. For each patient, a T2-weighted MRI scan is provided. The malignant group includes 81 hepatocellular carcinoma (HCC) and 13 intrahepatic cholangiocarcinoma (iCCA); the benign group includes 48 hepatocellular adenoma (HCA) and 44 FNH. The ground truth label, i.e., the phenotype of a lesion, was based on pathology. An exception are “typical” FNH [1], for which the ground truth was established radiologically. The differential diagnosis for each patient is provided, where patients have label 1 if the lesion was malignant, and label 0 if the lesion was benign. The phenotype of the lesions is also provided.

### 2.4. The GIST dataset

This dataset consists of 246 patients with either gastrointestinal stromal lesions (GISTs) (*N* = 125) or intra-abdominal tumors radiologically resembling GIST (non-GIST) (*N* = 122), as described in Starmans et al. [12]. One patient has two GISTs, thus the dataset in total contains 247 lesions. The non-GIST group consists of 22 schwannoma, 25 leiomyosarcoma, 25 leiomyoma, 25 esophageal or gastri junctional adenocarcinoma, and 25 lymphoma. For each patient, a contrast-enhanced venous phase CT scan is provided. The ground truth label, i.e., whether a lesion was a GIST or one of the non-GIST phenotypes, was confirmed by histology. The differential diagnosis for each patient is provided, where patients have label 1 if the lesion was a GIST, and label 0 if the lesion was a non-GIST. The subtype of the non-GIST lesions is also provided.

### 2.5. The CRLM dataset

This dataset consists of 77 patients with a total of 93 colorectal liver metastases (CRLM) with either a 100% desmoplastic histopathological growth patterns (HGP) [15] (*N* = 46) or 100% replacement HGP (*N* = 47), as described in Starmans et al. [13] ^2^. For each patient, a portal venous phase CT scan is provided. The ground truth label, i.e., whether a lesion had a desmoplastic or replacement HGP, was determined on hematoxylin and eosin stained tissue sections. The HGP type for each patient is provided, where patients have label 1 if the lesions had replacement HGP, and label 0 if the lesions had a desmoplastic HGP. As the HGP is assumed to be the same for all lesions of a subject, the ground truth is provided on subject level.

For each patient, for each lesion, segmentations by three clinicians (STUD1, PhD, RAD) and a Convolutional Neural Network (CNN) are available: e.g. *“segmentation_lesion1_STUD1*.*nii*.*gz”, “segmentation_lesion1_PhD*.*nii*.*gz”, “segmentation_lesion1_RAD*.*nii*.*gz”*, and *“segmentation_lesion1_CNN*.*nii*.*gz”*. Additionally, each lesion was segmented a second time by the first observer (STUD2), and is named e.g. *“segmentation_lesion1_STUD2*.*nii*.*gz”*. Note that 8 out of the 93 lesions (9%) were missed by the CNN, and thus do not include a CNN segmentation

### 2.6. The Melanoma dataset

This dataset consists of 169 lung metastases of 103 patients with *BRAF* mutated (*N* = 51) or *BRAF* wild type (*N* = 52) metastatic melanoma, as described in Angus et al. [14]. For each patient, a contrast-enhanced thoracic CT scan is provided. When multiple lesions were included, the corresponding segmentations are named *“segmentation_lesion1*.*nii*.*gz”, “segmentation_lesion2*.*nii*.*gz”*, and so on. The ground truth label, i.e., whether lesions from a patient were *BRAF* mutated or *BRAF* wild type, is provided, where patients have label 1 if the lesions were *BRAF* mutated, and label 0 if the lesions were *BRAF* wild type. As the *BRAF* mutation is assumed to be the same for all lesions of a subject, the ground truth is provided on subject level.

### 2.7. Acquisition protocol details

From the original DICOM files from the MRI and CT scans, the values of several tags were extracted to provide information on the used acquisition protocols, which for each scan are included in a *metadata*.*json* file.

For both MRI and CT scans, the following general acquisition protocol details from the following DICOM tags are included:

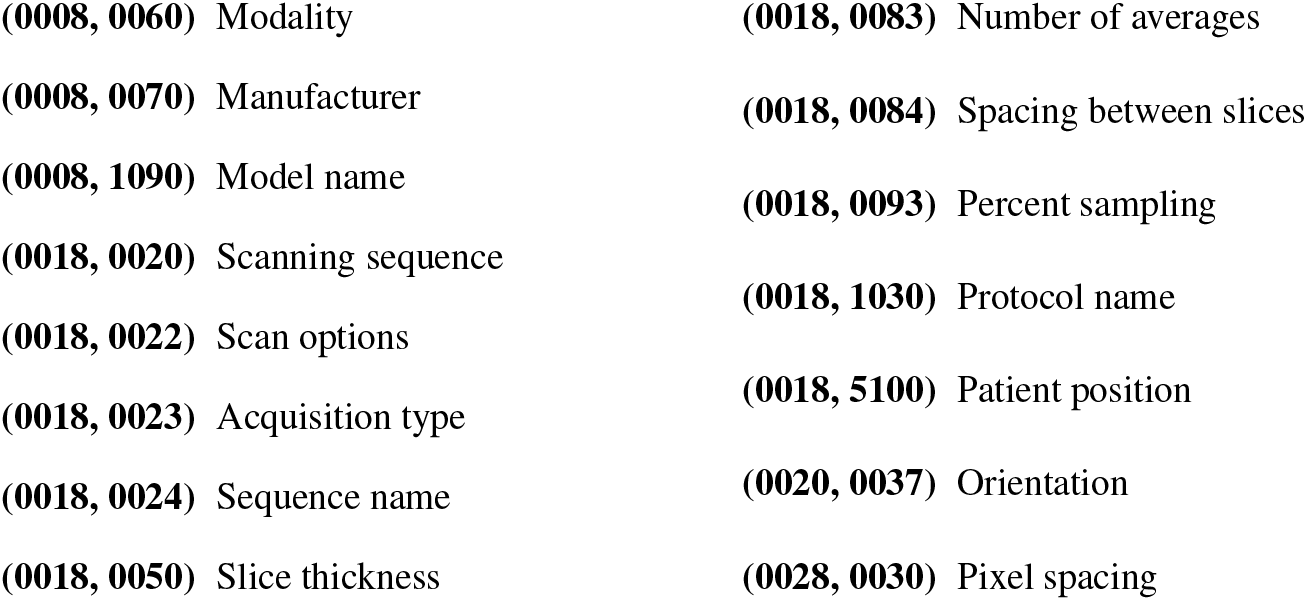

|  |  |
| --- | --- |
| **(0008, 0060)** Modality | **(0018, 0083)** Number of averages |
| **(0008, 0070)** Manufacturer | **(0018, 0084)** Spacing between slices |
| **(0008, 1090)** Model name | **(0018, 0093)** Percent sampling |
| **(0018, 0020)** Scanning sequence | **(0018, 1030)** Protocol name |
| **(0018, 0022)** Scan options | **(0018, 5100)** Patient position |
| **(0018, 0023)** Acquisition type | **(0020, 0037)** Orientation |
| **(0018, 0024)** Sequence name | **(0028, 0030)** Pixel spacing |
| **(0018, 0050)** Slice thickness |  |

For each MRI scan, the following specific acquisition protocol details from the following DICOM tags are additionally included:

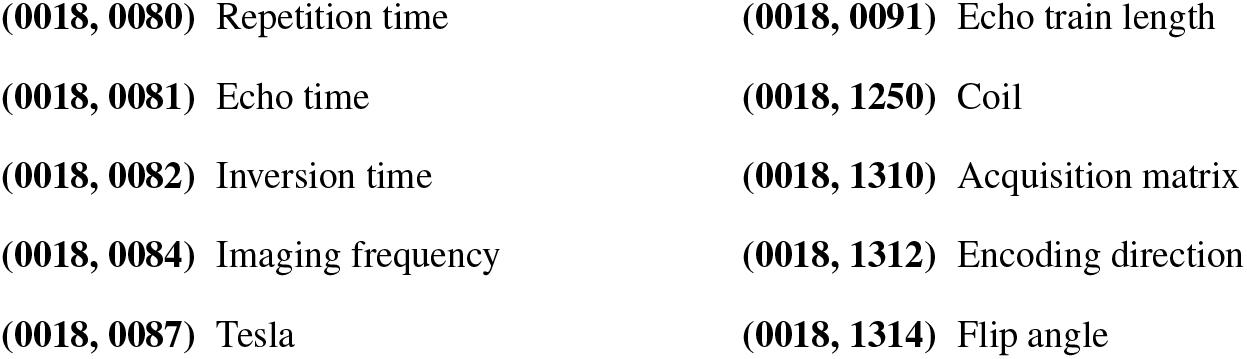

For each CT scan, the following specific acquisition protocol details from the following DICOM tags are additionally included:

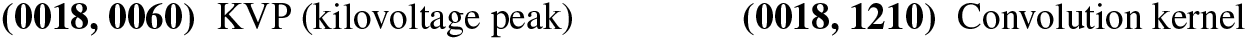

## 3. Experimental Design, Materials and Methods

### 3.1. The Lipo dataset

Patients that were either referred to/discussed at, or diagnosed/treated at the Erasmus MC Cancer Institute, Rotterdam, the Netherlands, between December 2009 and August 2018 with a pathologically confirmed diagnosis of lipoma or WDLPS were retrospectively included. Inclusion criteria were: a known *MDM2* amplification status tested by fluorescence *in situ* hybridization (FISH); and at least a T1-weighted MRI sequence available before treatment (if applicable).

The lipoma and WDLPS lesions were segmented semi-automatically on the T1-weighted MRI [16]. All images were segmented independently by either a medical masters student or a PhD candidate with an MD degree. Both were blinded to the type of lipomatous lesion. To validate segmentation accuracy, a sample set was verified by a musculoskeletal radiologist, specialized in soft-tissue sarcomas (4 years of experience). Semi-automatic results were always reviewed and manually corrected when necessary, to assure the result resembled manual segmentation.

### 3.2. The Desmoid dataset

Patients that were either referred to/discussed at, or diagnosed/treated at the Erasmus MC Cancer Institute, Rotterdam, the Netherlands, between 1990 and 2018 with histologically proven primary or recurrent DTF, or a malignant extremity STS, were retrospectively included. Inclusion criteria were: at least a T1-weighted MRI sequence available before treatment (if applicable); for the STS, a histologically proven primary myxofibrosarcoma, myxoid liposarcoma or leiomyosarcoma of the extremities.

The DTF and STS were all segmented semi-automatically on the T1-weighted MRI [16]. All images were segmented independently by either a medical masters student or a PhD candidate with an MD degree under supervision of a musculoskeletal radiologist (4 years of experience). Both were blinded to the type of lesion. Semi-automatic results were always reviewed and manually corrected when necessary, to assure the result resembled manual segmentation.

### 3.3. The Liver dataset

Patients that were either referred to/discussed at, or diagnosed/treated at the Erasmus MC Cancer Institute, Rotterdam, the Netherlands, between 2002 and 2018 with a primary solid liver lesion were retrospectively included. Inclusion criteria were: hepatocellular carcinoma (HCC), intrahepatic cholangiocarcinoma (iCCA), hepatocellular adenoma (HCA) or FNH; pathologically proven phenotype; and availability of a T2-weighted MRI scan. An exception to the pathologically proven phenotype was made for typical FNH, which are routinely not biopsied and diagnosed radiologically [17], as typical FNH imaging characteristics are 100% specific [18]. Exclusion criteria were: maximum diameter equal to or smaller than 3 cm; underlying liver disease; and significant imaging artefacts.

The lesions were all segmented semi-automatically on the T2-weighted MRI [16]. All images were segmented independently by one of two experienced abdominal radiologists (21 and 8 years of experience). Both were blinded to the type of lesion. Semi-automatic results were always reviewed and manually corrected when necessary, to assure the result resembled manual segmentation.

### 3.4. The GIST dataset

Patients that were either referred to/discussed at, or diagnosed/treated at the Erasmus MC Cancer Institute, Rotterdam, the Netherlands, between 2004 and 2017 with a histopathologically proven primary GIST or intra-abdominal tumors radiologically resembling GIST were retrospectively included. The inclusion criterion was availability of at least a contrast-enhanced venous-phase CT prior to treatment. The sample sizes of the non-GIST and the GIST cohort were matched. The non-GIST subtypes were balanced, i.e. a similar number of patients per subtype was randomly included.

The lesions were all segmented semi-automatically on the CT scan [16]. All images were segmented independently by either a medical masters student or a PhD candidate with an MD degree under supervision of a musculoskeletal radiologist (5 years of experience). Both were blinded to the type of lesion. Semi-automatic results were always reviewed and manually corrected when necessary, to assure the result resembled manual segmentation.

### 3.5. The CRLM dataset

Patients that were surgically treated at the Erasmus MC Cancer Institute, Rotterdam, the Netherlands, between 2003 and 2015 with CRLM were included. Inclusion criteria were: availability of at least a contrast-enhanced venous-phase CT prior to treatment; available hematoxylin and eosin stained tissue sections; either a 100% desmoplastic HGP or a 100% replacement HGP. Exclusion criteria were: recurrent CRLM or CRLM requiring two-staged resections; and treatment with preoperative chemotherapy, since chemotherapy may alter HGPs [15]. HGPs were scored on resection specimens according to the consensus guidelines by an expert pathologist (PV) [19].

The lesions were all segmented semi-automatically on the CT scan [16]. Lesion segmentation was performed by four observers: a medicine student with no relevant experience (STUD1), a PhD student (PhD) with limited experience, an expert abdominal radiologist (RAD), and an automatic CNN. The student segmented all lesions a second time (STUD2). All observers were blinded to the type of lesion. Semi-automatic results were always reviewed and manually corrected when necessary, to assure the result resembled manual segmentation.

The CNN used for the automatic segmentations was the Hybrid-Dense-UNet, which achieved state-of-the-art performance on the LITS liver tumor segmentation challenge and is open-source [20, 21]. The original CNN as trained on the LITS data was used. From the CNN lesion segmentations, only lesions that had histology were extracted, and the segmentations were saved per lesion.

### 3.6. The Melanoma dataset

Patients that were diagnosed with metastatic melanoma at the Erasmus MC Cancer Institute, Rotterdam, the Netherlands, between January 2012 and February 2018 were retrospectively included. Inclusion criteria were: known tumor *BRAF* mutation, diagnostic contrast-enhanced thoracic CT scan prior to commencement of any systemic therapy, and at least one lung metastasis of ≥ 10 mm evaluable according to Response Evaluation Criteria In Solid Tumors (RECIST) v1.1 [22]. Patients with *BRAF* mutations other than p.V600E were excluded. Formalin-fixed paraffin embedded material of the primary tumor and/ or metastasis was tested for *BRAF* (exon 15) using a polymerase chain reaction based assay or next generation sequencing as part of standard care.

Per patient, up to two lung lesions ≥ 10 mm were selected by a clinician supervised by an experienced chest radiologist and segmented semi-automatically on the CT scan [16]. In patients with >2 lung metastases of ≥10 mm, either the two largest or the two most easily distinguishable lesions were segmented (i.e., two separate lesions were preferred over two adjacent lesions). The clinician was blinded to the type of lesion. Semi-automatic results were always reviewed and manually corrected when necessary, to assure the result resembled manual segmentation.

## Data Availability

The data referred to in this manuscript is publicly available at https://xnat.bmia.nl/data/projects/worc. The code to download the data and reproduce the experiments from the radiomics study in which this data was presented can be found at https://github.com/MStarmans91/WORCDatabase.

https://github.com/MStarmans91/WORCDatabase

## 4. Ethics Statement

The study protocol for the collection of the WORC database conformed to the ethical guidelines of the 1975 Declaration of Helsinki. Approval by the local institutional review board of the Erasmus MC (Rotterdam, the Netherlands) was obtained for collection of the WORC database (MEC-2020-0961), and separately for the six included studies (Lipo: MEC-2016-339, Desmoid: MEC-2016-339, Liver: MEC-2017-1035, GIST: MEC-2017-1187, CRLM: MEC-2017-479, Melanoma: MEC-2019-0693). The need for informed consent was waived due to the use of anonymized, retrospective data.

## 5. Acknowledgments

The authors thank Laurens Groenendijk for his assistance in processing the data and in the anonimization procedures. Martijn P. A. Starmans acknowledges funding from the research program STRaTeGy with project numbers 14929 and 14930, which is (partly) financed by the Netherlands Organization for Scientific Research (NWO). Part of this study was financed by the Stichting Coolsingel (reference number 567), a Dutch non-profit foundation. This study is supported by EuCanImage (European Union’s Horizon 2020 research and innovation programme under grant agreement Nr. 952103).

## 6. CRediT Author Statement

M.P.A.S., M.J.M.T., M.V., D.J.G., C.V., S.S., F.E.B., L.A., A.A.M.v.d.V., R.L.M., M.G., J.J.V., W.J.N., and S.K. provided the conception and design of the study. M.P.A.S., M.J.M.T., M.V., G.A.P., D.J.G., C.V., S.S., G.J.L.H.v.L., F.E.B., F.E.J.A.W., B.G.K., L.A., A.A.M.v.d.V., A.R., A.E.O., M.R., M.D., R.d.M., J.IJ., R.L.M., P.B.V., M.G.T., and J.J.V. acquired the data. M.P.A.S., M.J.M.T., M.V., F.E.B., L.A., R.L.M., M.G.T. and S.K. analyzed and interpreted the data. M.P.A.S. created the software. M.P.A.S. and S.K. drafted the article. All authors read and approved the final manuscript.

## Footnotes

1 https://xnat.bmia.nl/data/projects/worc

2 Starmans et al. [13] reported a total of 76 patients, but the dataset did actually contain 77 patients.

